# Analysis of rare variants in 470,000 exome-sequenced UK Biobank participants implicates novel genes affecting risk of hypertension

**DOI:** 10.1101/2023.09.03.23294987

**Authors:** David Curtis

## Abstract

**Background:** A previous study of 200,000 exome-sequenced UK Biobank participants to test for association of rare coding variants with hypertension implicated two genes at exome-wide significance, *DNMT3A* and *FES*. A total of 42 genes had an uncorrected p value < 0.001. These results were followed up in a larger sample of 470,000 exome-sequenced participants.

**Methods and Results:** Weighted burden analysis of rare coding variants in a new sample of 97,050 cases and 172,263 controls was carried out for these 42 genes. Those showing evidence for association were then analysed in the combined sample of 167,127 cases and 302,691 controls. The association of *DNMT3A* and *FES* with hypertension was replicated in the new sample and they and the previously implicated gene *NPR1* were all exome-wide significant in the combined sample. Also exome-wide significant as risk genes for hypertension were *GUCY1A1*, *ASXL1* and *SMAD6*, while *GUCY1B1* had a nominal p value of < 0.0001. For two genes, *INPPL1* and *DBH*, rare coding variants predicted to impair gene function were protective against hypertension, again with exome-wide significance.

**Conclusions:** The findings offer new insights into biological risk factors for hypertension which could be the subject of further investigation. In particular, genetic variants predicted to impair the function of either membrane-bound guanylate cyclase, activated by natriuretic peptides, or soluble guanylate cyclase, activated by nitric oxide, increase risk of hypertension. Conversely, variants impairing the function of dopamine beta hydroxylase, responsible for the synthesis of norepinephrine, reduce hypertension risk.

This research has been conducted using the UK Biobank Resource.

## Introduction

An earlier analysis of the first release of exome sequence data for 200,000 UK Biobank participants identified two genes affecting risk of hypertension at exome-wide significance, *DNMT3A* and *FES* (Curtis, 2021). However since 20,384 genes were analysed the expected number of genes to be significant at *p* < 0.001 would be 20, whereas in fact 42 genes achieved this threshold, suggesting that some of these might represent true associations. Additionally, a number of these genes had functions which meant that they might plausibly be involved in affecting risk of hypertension.

Subsequently, UK Biobank has made available exome sequence data for 470,000 participants. The present study utilised data from the 270,000 participants not previously analysed and restricted attention to only the 42 genes previously achieving an SLP with absolute value greater than 3. For a subset of these genes which appeared to be of interest, data was analysed for all 470,000 participants to better assess the overall evidence implicating them and to gain insights into the categories of variant contributing to association.

It should be noted that most of this exome sequence data has also been used in two previous studies which tested for gene-wise associations with very large numbers of phenotypes and which included some hypertension-related phenotypes, so results from the present study need to be considered alongside these (Backman et al., 2021; Wang et al., 2021).

## Methods

UK Biobank participants are volunteers intended to be broadly representative of the UK population and are not selected on the basis of having any health condition. UK Biobank had obtained ethics approval from the North West Multi-centre Research Ethics Committee which covers the UK (approval number: 11/NW/0382) and had obtained informed consent from all participants. The UK Biobank approved an application for use of the data (ID 51119) and ethics approval for the analyses was obtained from the UCL Research Ethics Committee (11527/001). The UK Biobank Research Analysis Platform was used to access the Final Release Population level exome OQFE variants in PLINK format for 469,818 exomes which had been produced at the Regeneron Genetics Center using the protocols described here: https://dnanexus.gitbook.io/uk-biobank-rap/science-corner/whole-exome-sequencing-oqfe-protocol/protocol-for-processing-ukb-whole-exome-sequencing-data-sets (Backman et al., 2021). All variants were then annotated using the standard software packages VEP, PolyPhen and SIFT (Adzhubei et al., 2013; Kumar et al., 2009; McLaren et al., 2016). To obtain population principal components reflecting ancestry, version 2.0 of *plink* (https://www.cog-genomics.org/plink/2.0/) was run with the options *--maf 0.1 --pca 20 approx* (Chang et al., 2015; Galinsky et al., 2016).

Cases with hypertension were defined in exactly the same way as in the previous study, consisting of participants who met any of the following four criteria: a self-reported diagnosis recorded as hypertension or essential hypertension; self-reported taking medication for high blood pressure; reporting taking any of a list of named medications commonly used to treat high blood pressure (https://www.nhs.uk/conditions/high-blood-pressure-hypertension/); having an ICD10 diagnosis of essential hypertension, hypertensive heart disease or hypertensive renal disease in hospital records or as a cause of death (Curtis, 2021). Participants meeting any of these criteria were taken to be cases whereas all those meeting none of these criteria were taken to be controls. In the primary analyses to implicate specific genes attention was restricted to participants not included in the earlier study, consisting of 97,050 cases and 172,263 controls. For the subsequent analyses using the whole sample there were 167,127 cases and 302,691 controls.

The same analytic methods as had been used previously were applied, with the description repeated here for convenience. The SCOREASSOC program was used to carry out a weighted burden analysis to test whether, in each gene, sequence variants which were rarer and/or predicted to have more severe functional effects occurred more commonly in cases than controls. Attention was restricted to rare variants with minor allele frequency (MAF) <= 0.01 in both cases and controls. As previously described, variants were weighted by overall MAF so that variants with MAF=0.01 were given a weight of 1 while very rare variants with MAF close to zero were given a weight of 10 (Curtis, 2020). Variants were also weighted according to their functional annotation using the GENEVARASSOC program, which was used to generate the input files for weighted burden analysis by SCOREASSOC (Curtis, 2016, 2012). Variants predicted to cause complete loss of function (LOF) of the gene were assigned a weight of 100. Nonsynonymous variants were assigned a weight of 5 but if PolyPhen annotated them as possibly or probably damaging then 5 or 10 was added to this and if SIFT annotated them as deleterious then 20 was added. The full set of weights and categories is displayed in Table 1 of the previous study (Curtis, 2021). As described previously, the weight due to MAF and the weight due to functional annotation were multiplied together to provide an overall weight for each variant. Variants were excluded if there were more than 10% of genotypes missing in the controls or if the heterozygote count was smaller than both homozygote counts in the controls. If a subject was not genotyped for a variant then they were assigned the subject-wise average score for that variant. For each subject a gene-wise weighted burden score was derived as the sum of the variant-wise weights, each multiplied by the number of alleles of the variant which the given subject possessed.

**Table 1.** Genes with absolute value of SLP exceeding 3 (equivalent to p<0.001) for association of with hypertension in previous study showing the SLP obtained in the new sample. SLPs significant after correction for multiple testing are shown in bold. For all genes achieving an absolute value of SLP exceeding 2 and for *GUCY1B1*, the SLP obtained for combined sample is also shown.

| Symbol | SLP in original sample | Name | SLP in new sample | SLP in combined sample |
| --- | --- | --- | --- | --- |
| *DNMT3A* | 8.21 | DNA Methyltransferase 3 Alpha | **6.84** | 14.20 |
| *FES* | 6.10 | FES Proto-Oncogene, Tyrosine Kinase | **4.66** | 9.92 |
| *GUCY1A1* | 5.54 | Guanylate Cyclase 1 Soluble Subunit Alpha 1 | **5.06** | 9.13 |
| *NPR1* | 5.14 | Natriuretic Peptide Receptor 1 | 2.84 | 7.98 |
| *MAG* | 4.52 | Myelin Associated Glycoprotein | 0.12 |  |
| *SMAD6* | 4.10 | SMAD Family Member 6 | 2.43 | 6.02 |
| *GUCY1B1* | 3.93 | Guanylate Cyclase 1 Soluble Subunit Beta 1 | 1.88 | 4.95 |
| *SKIV2L* | 3.87 | Ski2 Like RNA Helicase | -0.33 |  |
| *ASXL1* | 3.75 | ASXL Transcriptional Regulator 1 | **5.34** | 8.35 |
| *MYO10* | 3.75 | Myosin X | -0.40 |  |
| *PTGER3* | 3.52 | Prostaglandin E Receptor 3 | -0.96 |  |
| *POLR3H* | 3.47 | RNA Polymerase III Subunit H | 0.45 |  |
| *IFT172* | 3.39 | Intraflagellar Transport 172 | -0.32 |  |
| *LITAF* | 3.37 | Lipopolysaccharide Induced TNF Factor | 0.54 |  |
| *CSK* | 3.37 | C-Terminal Src Kinase | 1.18 |  |
| *SNX27* | 3.34 | Sorting Nexin 27 | 0.10 |  |
| *TEK* | 3.31 | EK Receptor Tyrosine Kinase | 0.88 |  |
| *PHF21A* | 3.23 | PHD Finger Protein 21A | 0.90 |  |
| *LOC102723675* | 3.21 | Uncharacterized LOC102723675 | 0.12 |  |
| *DXO* | 3.14 | Decapping Exoribonuclease | 0.37 |  |
| *RHBDL3* | 3.12 | Rhomboid Like 3 | 0.08 |  |
| *ASB7* | 3.12 | Ankyrin Repeat And SOCS Box Containing 7 | 0.33 |  |
| *MPP2* | 3.11 | Membrane Palmitoylated Protein 2 | 0.38 |  |
| *ZNF549* | 3.10 | Zinc Finger Protein 549 | 0.55 |  |
| *MAP3K14-AS1* | 3.08 | MAP3K14 Antisense RNA 1 | 1.69 |  |
| *ZNF600* | -3.01 | Zinc Finger Protein 600 | -0.28 |  |
| *SIRT2* | -3.05 | Sirtuin 2 | -0.67 |  |
| *PARGP1* | -3.05 | Poly(ADP-Ribose) Glycohydrolase Pseudogene 1 | 1.16 |  |
| *ZNF226* | -3.13 | Zinc Finger Protein 226 | 0.15 |  |
| *LOC105375153* | -3.18 | Uncharacterized LOC105375153 | 0.69 |  |
| *INPPL1* | -3.19 | Inositol Polyphosphate Phosphatase Like 1 | **-4.02** | -7.09 |
| *FNDC8* | -3.23 | Fibronectin Type III Domain Containing 8 | -0.43 |  |
| *KRT82* | -3.26 | Keratin 82 | -0.47 |  |
| *GOLGA4* | -3.36 | Golgin A4 | -0.51 |  |
| *DBH* | -3.40 | Dopamine Beta-Hydroxylase | **-5.61** | -9.71 |
| *ELOF1* | -3.59 | Elongation Factor 1 Homolog | 0.08 |  |
| *AGTR1* | -3.77 | Angiotensin II Receptor Type 1 | -0.76 |  |
| *SIPA1* | -3.81 | Signal-Induced Proliferation-Associated 1 | -1.41 |  |
| *ZYX* | -3.83 | Zyxin | -0.31 |  |
| *ZFAT* | -4.00 | Zinc Finger And AT-Hook Domain Containing | -0.34 |  |
| *OSTF1* | -4.05 | Osteoclast Stimulating Factor 1 | 0.17 |  |
| *PREP* | -5.03 | Prolyl Endopeptidase | -0.11 |  |

Analyses were restricted to the 42 genes significant at p < 0.001 in the previous study. For each gene, logistic regression analysis was carried out with hypertension as the dependent variable including the first 20 population principal components and sex as covariates and a likelihood ratio test was performed comparing the likelihoods of the models with and without the gene-wise burden score. This is a test for association between the gene-wise burden score and caseness and the statistical significance was summarised as a signed log p value (SLP), which is the log base 10 of the p value given a positive sign if the score is higher in cases and negative if it is higher in controls. Since only 42 genes were analysed, after correcting for multiple testing a gene could be declared statistically significant if it achieved an SLP with absolute value greater than −log10(0.05/42) = 2.9 using the new samples.

Follow-up analyses were performed on all genes individually achieving an absolute value for the SLP of 2 (equivalent to p = 0.01) and also *GUCY1B1*. For this subset of genes the weighted burden analysis described above was carried out using the whole sample of 167,127 cases and 302,691 controls. Additionally, for each subject a count was obtained of the number of variants they carried falling into particular broad categories, such as LOF, protein altering, etc. The full list of these categories is shown in Supplementary Table 1. These counts were entered into a multiple logistic regression analysis with hypertension as the dependent variable and again including sex and 20 principal components as covariates in order to elucidate the contribution of different types of variant to the overall evidence for association. The odds ratios (ORs) associated with each category were estimated along with their standard errors and the Wald statistic was used to obtain a p value. This p value was converted to an SLP, again with the sign being positive if the OR was greater than 1, indicating that variants in that category tended to increase risk.

Data manipulation and statistical analyses were performed using GENEVARASSOC, SCOREASSOC and R (R Core Team, 2014).

## Results

Table 1 shows the results of the primary analysis. The two genes which were exome-wide significant in the previous study, *DNMT3A* and *FES*, again show convincing evidence of association in the new sample with SLPs of 6.84 and 4.66 respectively. Additionally, four genes which were not exome-wide significant in the previous sample produce results which are significant after correction for multiple testing in the new sample: *GUCY1A1* (SLP = 5.06), *ASXL1* (SLP = 5.34), *INPPL1* (SLP = −4.02) and *DBH* (SLP = −5.61). These genes were taken forward for secondary analysis in the combined sample and it is noteworthy that, as also shown in Table 1, all six of them then produced SLPs with absolute value exceeding the threshold of 5.61 to be regarded as exome wide significant. All other genes producing an SLP with absolute value exceeding 2 were also analysed in the combined sample, along with *GUCY1B1* which produced an SLP of only 1.88 but which codes for a subunit of the same guanylate cyclase as *GUCY1A1*. Two of these genes produced SLPs in the combined sample exceeding the criterion for exome wide significance, *NPR1* (SLP = 7.98) and *SMAD6* (SLP = 6.02). In the combined sample *GUCY1B1* produced an SLP of 4.95.

In order to gain insights into the effects of different categories of variant within these genes of interest, counts for variants of each category in each subject were entered into multiple logistic regression analysis along with sex and 20 principal components as covariates. Table 2 shows results for all genes and categories where the category was significant at p < 0.05, with an absolute value of SLP > 1.3. Additionally, for any gene in which the SIFT or PolyPhen annotations were significant the results for the protein altering category are also shown, because the annotations only apply to protein altering variants. (Full results for all genes and all categories are shown in Supplementary Table 2.)

**Table 2.**
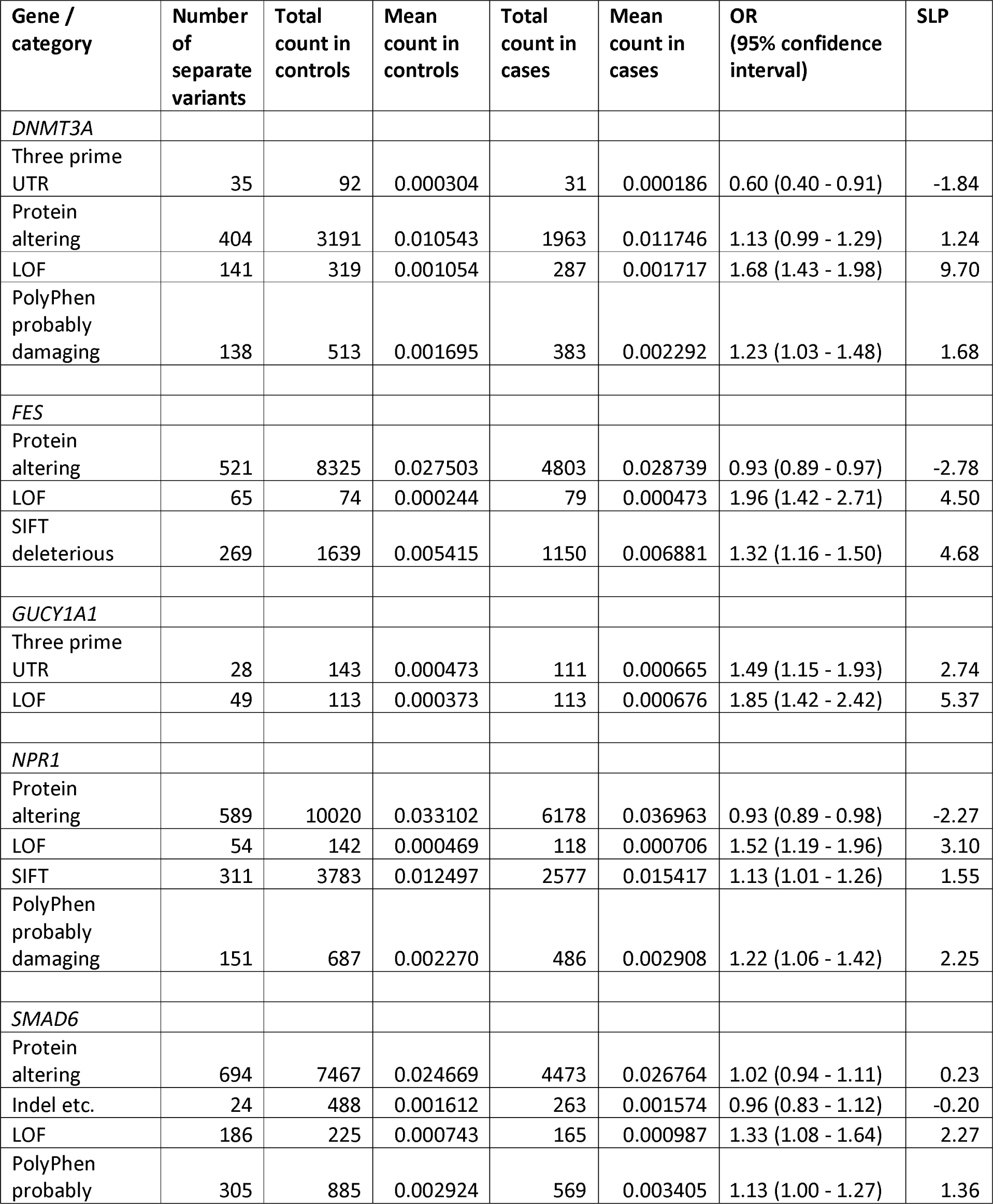
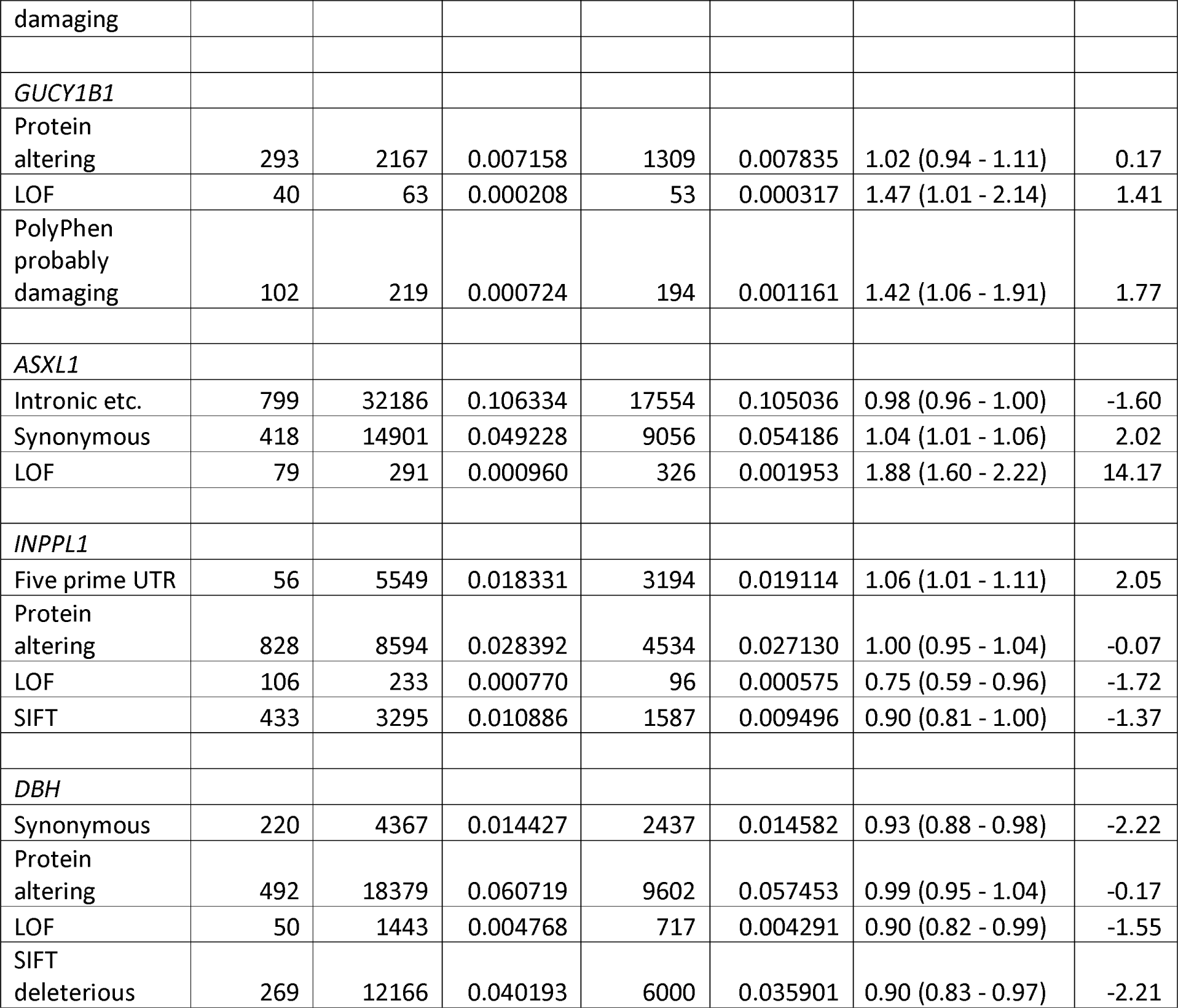
The table shows results obtained from multiple logistic regression analyses including all variant categories along with sex and 20 principal components as covariates . Results are shown for each category of variant for which the category-wise results are nominally significant at p < 0.05 (absolute value of SLP > 1.3). Where the results of SIFT or PolyPhen were significant, the results are also shown for all protein altering variants.

A number of findings are of interest. For every gene, LOF variants produce the largest effect. For the genes in which impaired functioning increases hypertension risk, the OR for a LOF variant ranges from 1.33 for *SMAD6* to 1.96 for *FES*. For the genes in which impaired function is protective against hypertension, the OR for a LOF variant is 0.75 for *INPPL1* and 090 for *DBH*. Although LOF variants are associated with the largest effect size they are also very rare and for most genes the mean number of LOF variants carried per subject is fewer than 0.001. By contrast, other categories of variant tend to be commoner but to be associated with smaller effect sizes. For example, in *DNMT3A* a LOF variant has an OR of 1.68 but a non-synonymous variant annotated as probably damaging by SIFT has an OR of 1.13 x 1.23 = 1.39. The table shows that there is a lack of consistency across genes in terms of which categories of variant show evidence of association. It is generally the case that variants in categories associated with an effect have low cumulative frequencies but it is perhaps noteworthy that non-synonymous variants in *DBH* which are annotated as deleterious by SIFT have OR of 0.9 and the average number carried per subject is greater than 0.035. This means that about 1 person in 30 carries a *DBH* variant of a category which on average slightly reduces their risk of hypertension.

## Discussion

This study confirms that rare coding variants which impair the function of the two genes identified in the earlier study, *DNMT3A* and *FES*, do indeed increase the risk of hypertension. Coding variants in *NPR1* have previously been reported to be associated with hypertension risk and this gene is also exome-wide significant in the combined sample (Liu et al., 2016; Vandenwijngaert et al., 2019). Four genes are newly implicated at conventional levels of significance, whether one corrects for the multiple testing of 42 genes in this study or the 20,384 originally considered, these being *GUCY1A1*, *ASXL1*, *INPPL1* and *DBH*. Variants damaging the function of *GUCY1A1* or *ASXL1* increase hypertension risk while variants in *INPPL1* and *DBH* are protective. For two other genes the results are somewhat more ambiguous. *SMAD6* failed to achieve statistical significance in the primary analysis of the new sample and barely reaches the criterion of exome-wide significance in the combined sample. *GUCY1B1* was not statistically significant in the primary analysis and nor is it exome-wide significant overall but it is such a biologically plausible candidate that it does not seem sensible to simply ignore it.

As mentioned above, this dataset has been used for previous two studies testing for association between exome sequence variance with a very large number of phenotypes, which for convenience we can refer to as the Regeneron and AstraZeneca studies (Backman et al., 2021; Wang et al., 2021). The Regeneron study carried out a variety of single variant and gene-wise burden tests on 3,994 health-related traits to produce a total of about 2.3 billion tests, yielding a critical p value of 2.18[]×[]10 (corresponding to SLP = 10.66)) and reported 8,865 significant associations which are presented in their Supplementary Data 2 (Backman et al., 2021). This contains no gene-wise associations for any of these genes with any hypertension related phenotype but does include one single variant association of a nonsynonymous variant in *DBH*, 9:133636634:G:C, with reduced diastolic blood pressure at 5.48 x 10^14^ (SLP = −13.26). For the AstraZeneca study, all gene-wise and variant-wise associations with 17,361 binary and 1,419 quantitative phenotypes are reported on the AstraZeneca PheWAS Portal at https://azphewas.com/ (Wang et al., 2021). This was accessed to find the most significant p value for any analysis of each of these genes with the phenotype "ICD10 I10 Essential (primary) hypertension" and Table 3 shows the results obtained compared with those for the current study. It can be seen that for each gene except *ASXL1* the AstraZeneca results provide less support for association than the present study.

**Table 3.** Comparison of results from current study to those reported for the AstraZeneca study. The results for the AstraZeneca study are displayed as the equivalent SLP for the most significant result reported for that gene with the phenotype ICD10 Essential (primary) hypertension.

| Gene | SLP for combined sample in current study | SLP for AstraZeneca study |
| --- | --- | --- |
| *DNMT3A* | 14.20 | 8.19 |
| *FES* | 9.92 | 5.04 |
| *GUCY1A1* | 9.13 | 6.61 |
| *NPR1* | 7.98 | 4.12 |
| *SMAD6* | 6.02 | 2.25 |
| *GUCY1B1* | 4.95 | 1.70 |
| *ASXL1* | 8.35 | 10.17 |
| *INPPL1* | -7.09 | -3.45 |
| *DBH* | -9.71 | -4.12 |

There are a number of differences between the approach used here and that used in the Regeneron and AstraZeneca studies which might account for differences in the results obtained. One is that the Regeneron study analyses are based on []430,998 participants with European ancestry and the AstraZeneca study used 394,695 exomes which were of high quality from participants who were predominantly unrelated and of European ancestry while results for the current study are based on all 469,818 exomes made available in the final release. Another difference is that the other studies used multiple phenotypes and multiple methods of single variant and gene-based analyses, necessitating a rigorous correction for multiple testing, meaning that some results potentially of interest might not be identified. Another difference is that the present study uses a definition of caseness which combines information from a number of sources rather than using a single measure. Finally, the present study implements a weighted burden analysis in which all variants are analysed together but variants which are rare and/or predicted to have more serious consequences are accorded higher weights. The other studies implemented burden analyses using different sets of variant category. Thus, one analysis might only include a small number of LOF variants and another might include LOF variants along with a much larger number of nonsynonymous variants. However, without weighting the LOF variants more strongly there could be a risk that their signal would be swamped out by the other variants. Finally, it should be clearly stated that although the present study arguably highlights some genes of interest more effectively than the other studies it also misses associations which were picked up by the others. The Regeneron study did report a risk-lowering association of *SLC9A3R2* with hypertension and measured blood pressure (SLP = −18.70) while the AstraZeneca study found a significant association of *PKD1* with hypertension (SLP = 12.28) but neither of these genes were picked up in the current investigation.

Another report has recently been published which used the same dataset to test for association between LOF variants and systolic or diastolic blood pressure (Lecluze and Lettre, 2023). They report single variant associations with ten genes (*ANKDD1B*, *ENPEP*, *PNCK*, *BTN3A2*, *C1orf145*, *CASP9*, *DBH*, *KIAA1161*, *OR4X1* and *TMC3*) and associations with burden of LOF variants for five genes (*TTN*, *NOS3*, *FES*, *SMAD6* and *COL21A1*). Thus, the only gene-wise results which overlap with this study are for *FES* and *SMAD6*. The variant association they report for *DBH* is with the splice site variant 9:133636712:T:C but the minus log10 p value they report for this is only 6.79, which arguably should not be regarded as statistically significant given that 515,198 LOF variants were tested. Their approach differed from the current study in that they considered only LOF variants, that they carried out single variant analyses as well as gene-based burden tests and that they used measured blood pressure rather than a clinical hypertension phenotype.

For at least some of the implicated genes there are plausible biological mechanisms which might contribute to the observed association. As discussed previously, DNMT3A methylates DNA conditional on the associated H3K4 residue being unmethylated hence has effects on gene expression and in mice, *Dnmt3a* knockdown has been shown to lead to reduced methylation of the gene for angiotensin receptor type 1a, *Agtr1a*, leading to increased *Agtr1a* expression and salt-induced hypertension (33). It has been reported that in human children and adolescents, DNMT3A levels are positively associated with obesity and with diastolic blood pressure (Salah et al., 2022). However the present study indicates that impaired functioning of *DNMT3A* increases rather than decreases the risk of hypertension. Another implicated gene, *ASXL1*, is also a transcriptional regulator and is involved in the deubiquitination of histone H2A lysine 119 which is frequently mutated in cancers, but the mechanism whereby impaired function might increase hypertension risk remains to be elucidated (Thomas et al., 2023). In Bohring-Opitz syndrome, characterised by a prominent metopic suture, hypertelorism, exophthalmos, cleft lip and palate, limb anomalies, as well as difficulty feeding with severe developmental delays, almost 50% have *de novo* LOF variants in *ASXL1* (Dangiolo et al., 2015; Hoischen et al., 2011). However the finding that over 600 UK Biobank participants carry *ASXL1* LOF variants shows that Bohring-Opitz syndrome is by no means an inevitable consequence and further work could address whether specific subsets of LOF variants, perhaps affecting particular transcripts, are relevant.

Perhaps the genes for which a biological mechanism is most obvious are *NPR1*, *GUCY1A1* and *GUCY1B1*. *NPR1* codes for a membrane bound guanylate cyclase which acts as a receptor for natriuretic peptides and variants in this gene associated with blood pressure have been shown to modulate guanylate cyclase activity (Vandenwijngaert et al., 2019). *GUCY1A1* and *GUCY1B1* code for subunits of a soluble guanylate cyclase which responds to nitric oxide signalling and has a well established role in blood pressure regulation (Buys and Sips, 2014). Guanylate cyclase produces cyclic GMP which acts as an intracellular messenger to mediate responses such as vasodilation and the results reported here further reinforce the notion that impaired guanylate cyclase activity is a risk factor for hypertension.

Another gene for which the role in hypertension susceptibility is fairly obvious is *DBH*. Dopamine beta hydroxylase catalyzes the oxidative hydroxylation of dopamine to norepinephrine in the adrenal medulla and the synaptic vesicles of postganglionic sympathetic neurons, and variants affecting both copies of the gene can act recessively to cause norepinephrine deficiency syndrome, characterised by orthostatic hypotension among other symptoms (Kim et al., 2002). The present results demonstrate that variants affecting a single copy of the gene can be protective against hypertension, presumably by leading to a more modest reduction in norepinephrine levels. In animal studies, inhibition of dopamine beta hydroxylase using nepicastat reduces behaviours associated with use of cocaine and morphine (Frankowska et al., 2021; Schroeder et al., 2013). Nepicastat, was trialled as a treatment for post-traumatic stress disorder and for cocaine dependence but although both trials are completed no results have been published (https://clinicaltrials.gov/study/NCT00641511, https://clinicaltrials.gov/study/NCT00656357). It is unclear whether dopamine beta hydroxylase might be a suitable drug target for the management of hypertension. In order to avoid central effects, it might be productive to investigate therapeutic agents which did not cross the blood brain barrier.

As for *ASXL1*, it is not immediately clear by what mechanisms *FES*, *SMAD6* or *INPPL1* might impact risk of hypertension and if the results of the present study appear convincing then this could act as a stimulus to carrying out further research into the function of these genes.

Overall, this study implicates a small number of genes in which variants predicted to impair gene function can increase or decrease risk of hypertension. The cumulative allele frequency of these variants is rare and hence these results do not have clinical or public health implications. Rather, the findings may focus attention on particular pathways important to moderating the risk of hypertension and this might ultimately lead to the identification of novel drug targets and treatment strategies.

## Data availability

The raw data is available on application to UK Biobank. Detailed results with variant counts cannot be made available because they might be used for subject identification. Scripts and relevant derived variables will be deposited in UK Biobank. Software and scripts used to carry out the analyses are also available at https://github.com/davenomiddlenamecurtis.

## Data Availability

https://www.ukbiobank.ac.uk/

## Acknowledgments

This research has been conducted using the UK Biobank Resource. The author wishes to acknowledge the staff supporting the High Performance Computing Cluster, Computer Science Department, University College London. This work was carried out in part using resources provided by BBSRC equipment grant BB/R01356X/1. The author wishes to thank the participants who volunteered for the UK Biobank project.

## Ethics statement

UK Biobank had obtained ethics approval from the North West Multi-centre Research Ethics Committee which covers the UK (approval number: 11/NW/0382) and had obtained informed consent from all participants. The UK Biobank approved an application for use of the data (ID 51119) and ethics approval for the analyses was obtained from the UCL Research Ethics Committee (11527/001).

## Author contributions

DC carried out the analyses and prepared the manuscript.

---

**Supplementary Table 1.** The table shows the broad categories used for variant category specific analyses along with the annotations produced by VEP which were grouped into each category.

| Category | VEP annotation |
| --- | --- |
| Intronic etc. | feature_truncation, regulatory_region_variant, feature_elongation, regulatory_region_amplification, regulatory_region_ablation, TF_binding_site_variant, TFBS_amplification, TFBS_ablation, downstream_gene_variant, upstream_gene_variant, non_coding_transcript_variant, NMD_transcript_variant, intron_variant, non_coding_transcript_exon_variant |
| Five prime UTR | 5_prime_UTR_variant |
| Synonymous | synonymous_variant |
| Splice region | splice_region_variant |
| Three prime UTR | 3_prime_UTR_variant |
| Protein altering | protein_altering_variant, missense_variant |
| Indel etc. | inframe_deletion, inframe_insertion, transcript_amplification |
| LOF | frameshift_variant, stop_gained, transcript_ablation, splice_donor_variant, splice_acceptor_variant |
| SIFT deleterious | deleterious |
| PolyPhen possibly damaging | possibly_damaging |
| PolyPhen probably damaging | probably_damaging |

**Supplementary Table 2.**
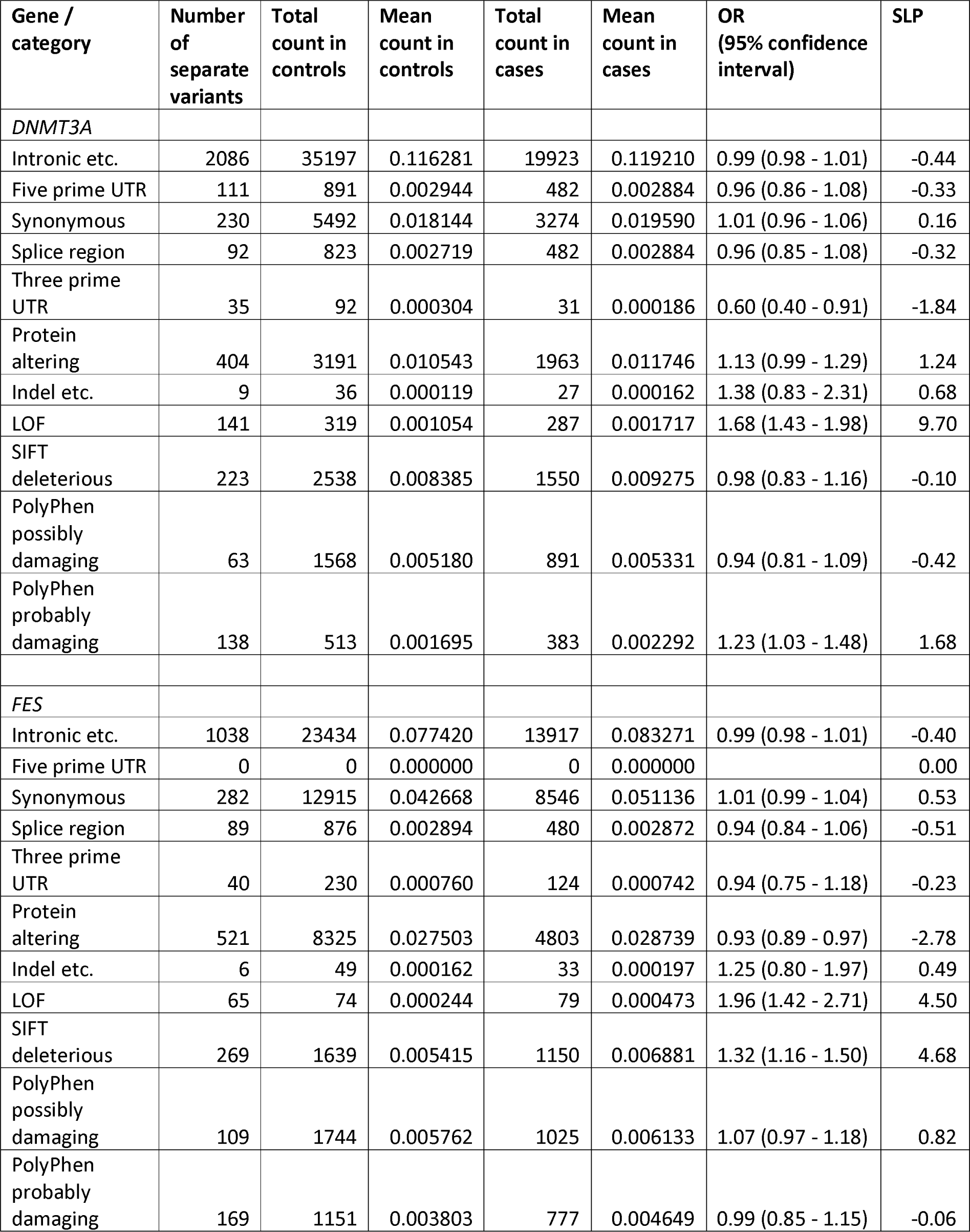
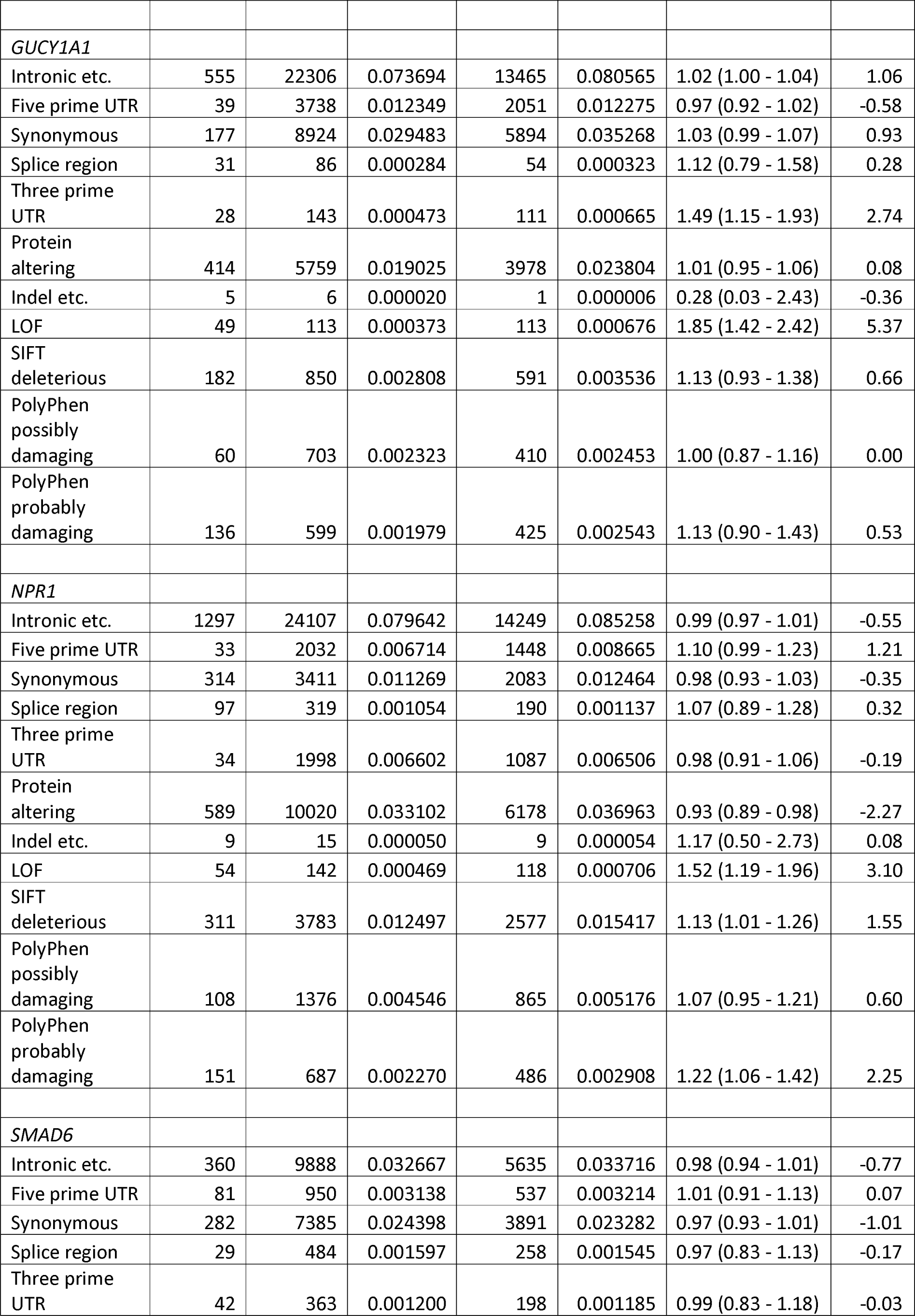
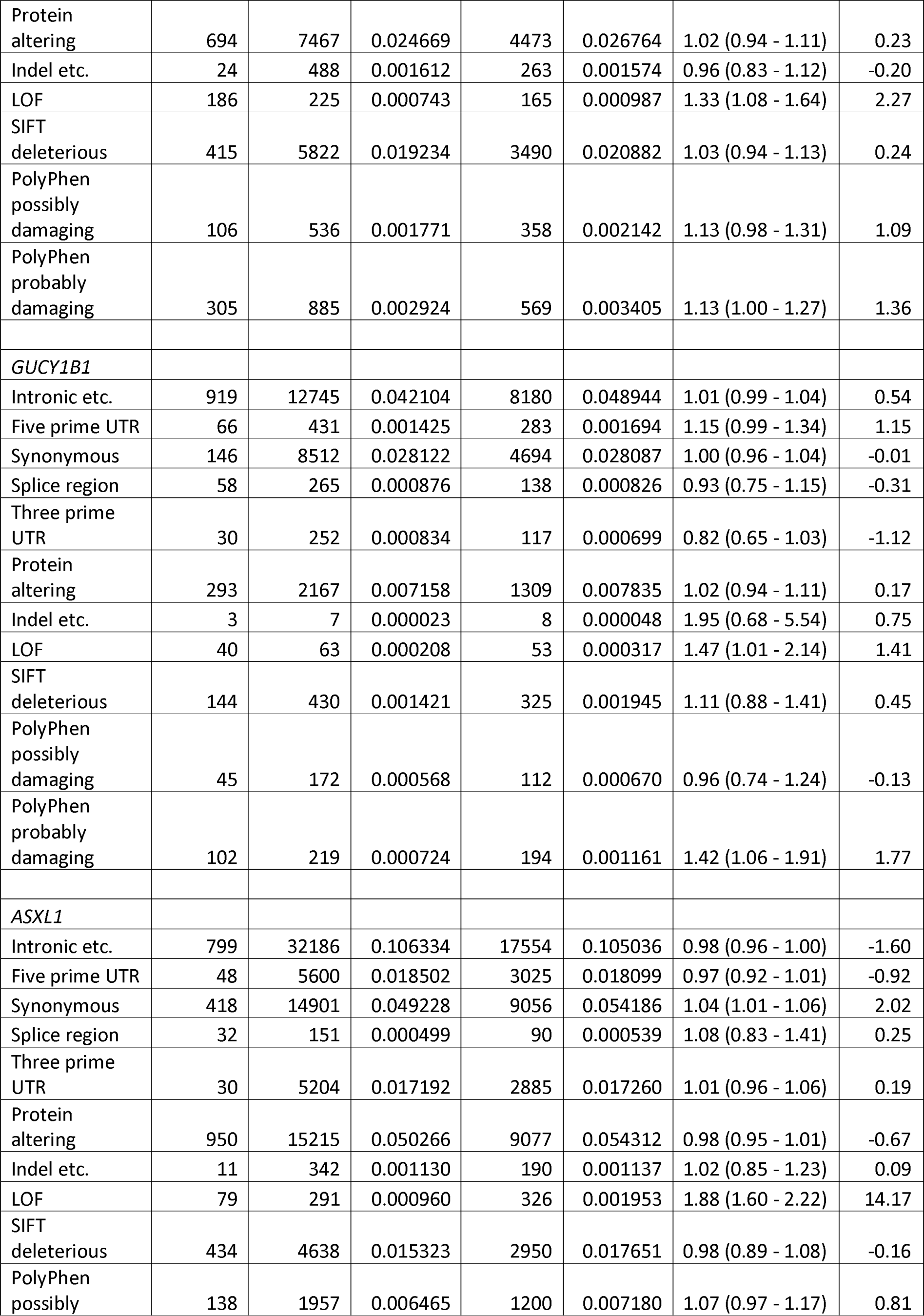
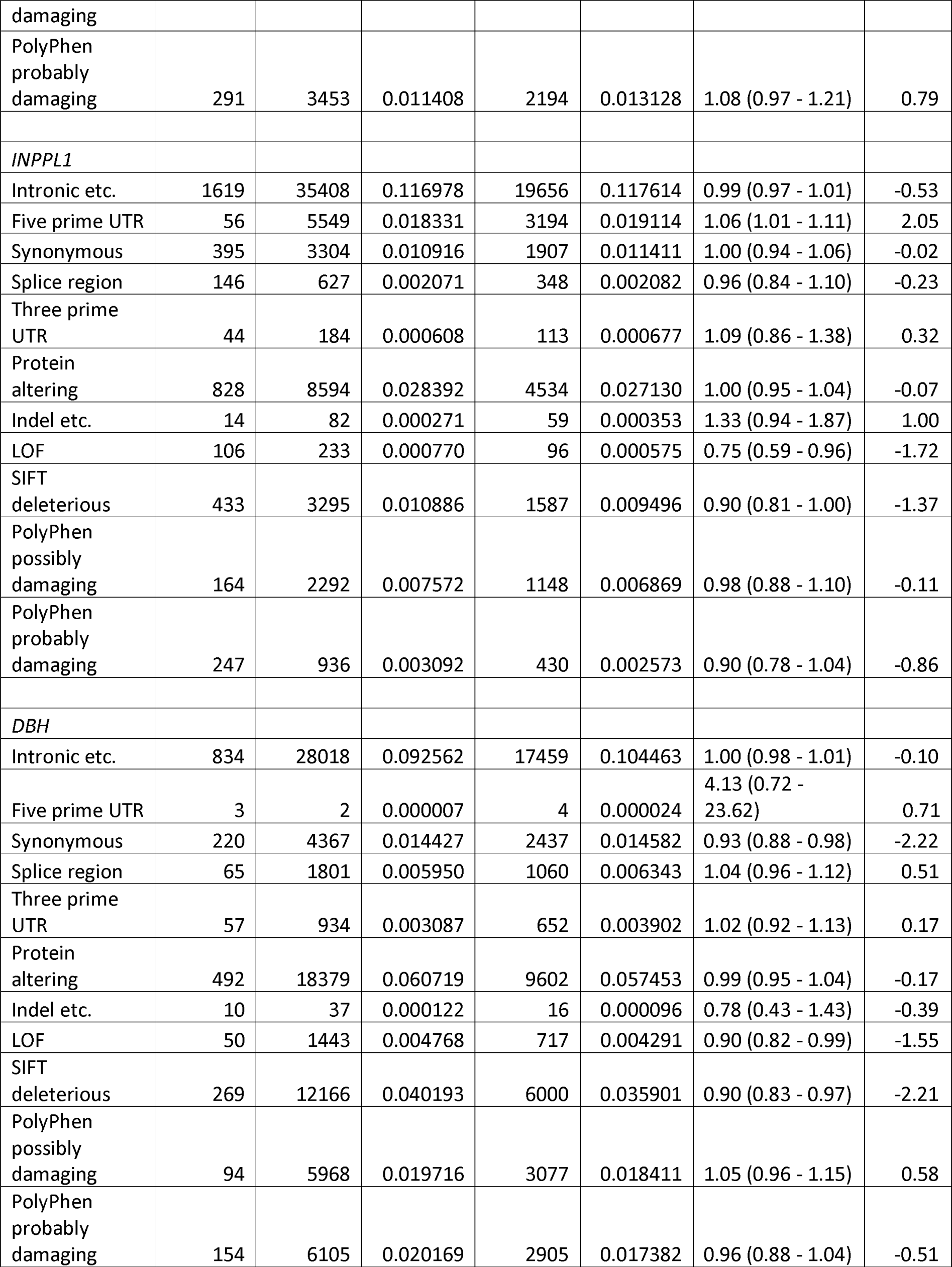
The table shows results for every category of variant in each gene of interest for a multiple logistic regression analysis including all variant categories along with sex and 20 principal components as covariates.

